# Clinical and microbiological assessments of COVID-19 in healthcare workers: a prospective longitudinal study

**DOI:** 10.1101/2020.11.04.20225862

**Authors:** Antonin Bal, Karen Brengel-Pesce, Alexandre Gaymard, Grégory Quéromès, Nicolas Guibert, Emile Frobert, Maude Bouscambert, Mary-Anne Trabaud, Florence Allantaz-Frager, Guy Oriol, Valérie Cheynet, Constance d’Aubarede, Amélie Massardier-Pilonchery, Marlyse Buisson, Julien Lupo, Bruno Pozzetto, Pascal Poignard, Bruno Lina, Jean-Baptiste Fassier, Florence Morfin-Sherpa, Sophie Trouillet-Assant, on behalf of COVID SER STUDY GROUP

**Affiliations:** Laboratoire de Virologie, Institut des Agents Infectieux, Laboratoire associé au Centre National de Référence des virus des infections respiratoires, Hospices Civils de Lyon, Lyon, France; CIRI, Centre International de Recherche en Infectiologie, Team VirPath, Univ Lyon, Inserm, U1111, Université Claude Bernard Lyon 1, CNRS, UMR5308, ENS de Lyon, F-69007, Lyon, France; Joint Research Unit Hospices Civils de Lyon-bioMérieux, Lyon Sud Hospital, Pierre-Bénite, France; Lyon University, Université Claude Bernard Lyon1, Ifsttar, UMRESTTE, UMR T_9405, 8 avenue Rockefeller Lyon, France; Occupational Health and Medicine Department, Hospices Civils de Lyon, Lyon, France; biomérieux; Institut de Biologie Structurale, Université Grenoble Alpes, CEA, CNRS and Centre Hospitalier Universitaire Grenoble Alpes, Grenoble, France; GIMAP EA 3064 (Groupe Immunité des Muqueuses et Agents Pathogènes), Université Jean Monnet, Lyon University, Saint-Etienne, France; Laboratory of Infectious Agents and Hygiene, University Hospital of Saint-Etienne, Saint-Etienne, France

**Keywords:** COVID-19, SARS-CoV-2, Health-care workers, viral load, viral culture, RT-PCR, neutralizing antibody

## Abstract

**Background:** A comprehensive assessment of COVID-19 in healthcare workers (HCWs) including the investigation of viral shedding duration is critical.

**Methods:** A longitudinal study including 319 HCWs was conducted. After SARS-CoV-2 screening with RT-PCR assay, other respiratory pathogens were tested with a multiplex molecular panel. For SARS-CoV-2 positive HCWs, the normalized viral load was determined weekly; viral culture and virus neutralization assays were also performed. For 190 HCWs tested negative, SARS-CoV-2 serological testing was performed one month after the inclusion.

**Findings:** Of the 319 HCWs included, 67 (21.0%) were tested positive for SARS-CoV-2; two of them developed severe COVID-19. The proportion of smell and taste dysfunction was significantly higher in SARS-CoV-2 positive HCWs than in negative ones (38.8% vs 9.5% and 37.3% vs 10.7%, respectively, p<0.001). Of the 67 positive patients, 9.1% were tested positive for at least another respiratory pathogen (*vs* 19.5%, p=0.07). The proportion of HCWs with a viral load > 5.0 log_10_ cp/ml (Ct value <25) was less than 15% at 8 days after symptom onset; 12% of them were still positive after 40 days (Ct >37). More than 90% of culturable virus had a viral load > 4.5 log_10_ cp/ml (Ct < 26) and were collected within 10 days after symptom onset. From HCWs tested negative, 6/190 (3.2%) exhibited seroconversion for IgG antibodies.

**Interpretation:** Our data suggest that the determination of normalized viral load (or its estimation through Ct values) can be useful for discontinuing isolation of HCWs and facilitating their safe return to work. HCWs presenting mild COVID-19 are unlikely infectious 10 days after symptom onset.

**Funding:** Fondation des Hospices Civils de Lyon. bioMérieux provided diagnostic kits.

## Introduction

Since the beginning of the SARS-CoV-2 pandemic in December 2019, healthcare workers (HCWs) from all over the world have been on the front line for the management of COVID-19 patients. Due to close, repeated, and prolonged contact with SARS-CoV-2 infected patients, HCWs have been a privileged target of the COVID-19 pandemic [1–3]. Similar to the rest of the population, the clinical spectrum of SARS-CoV-2 infections reported in HCWs encompassed asymptomatic, mild but also severe and fatal infections [4–7]. The early detection of SARS-CoV-2 infected HCWs is crucial to reduce the risk of nosocomial transmission, which is associated with an important mortality when occurring between at-risk patients including elderly people [6,8,9]. However, medical settings may not have enough resources to keep infected HCWs on leave for a long time due to HCW shortage. Defining the duration of infectivity of HCWs is therefore of paramount importance for their appropriate management, which becomes crucial to face the present intensive recirculation of the virus in the Northern hemisphere [10,11].

The diagnosis of SARS-CoV-2 infection is mainly based on RT-PCR performed on naso-pharyngeal swabs (NPS). The virus can be detected about 2 to 3 days before the onset of symptoms and the viral RNA excretion can last up to several weeks depending on the immune competence, the patient age as well as the severity of the disease [12–17]. As frequently observed for other viral infections, the SARS-CoV-2 viral RNA can be detected beyond resolution of symptoms, after seroconversion, and without any detectable infectious virus in clinical samples [18–21]. To assess the duration of infectivity in COVID-19 patients, no standard, rapid, and reliable method is available, and so viral isolation in cell culture remains the most appropriate approach despite its fastidiousness [22]. In previous reports, SARS-CoV-2 isolation could be performed up to 10 days after symptom onset in mild patients [18,23–25] and up to 22 days after the first positive result in severe patients [20,25–27]. To enhance clinical management of SARS-CoV-2 infection in HCWs, a virological investigation including quantitative RT-PCR, viral culture as well as the determination of neutralizing antibodies titers over the course of the disease is needed. With this aim, we performed a comprehensive assessment of COVID-19 in a longitudinal cohort study of 319 front-line HCWs enrolled during the first wave of the pandemic.

## Methods

### Study design

A prospective longitudinal cohort study was conducted at the university hospital of Lyon, France (*Hospices Civils de Lyon*, HCL) [28] in HCWs with symptoms suggesting a SARS-CoV-2 infection. HCWs with a previous positive SARS-CoV-2 RT-PCR test were excluded. Clinical and microbiological data were recorded for all included HCWs. Patients with a positive RT-PCR result at inclusion (V1) came back during 6 consecutive weeks for SARS-CoV-2 (V2-V7) quantitative RT-PCR on NPS and blood sampling at each time point. HCWs with negative SARS-CoV-2 PCR at inclusion came back one month later (V5) for SARS-CoV-2 serology testing (Figure 1).

**Figure 1.**
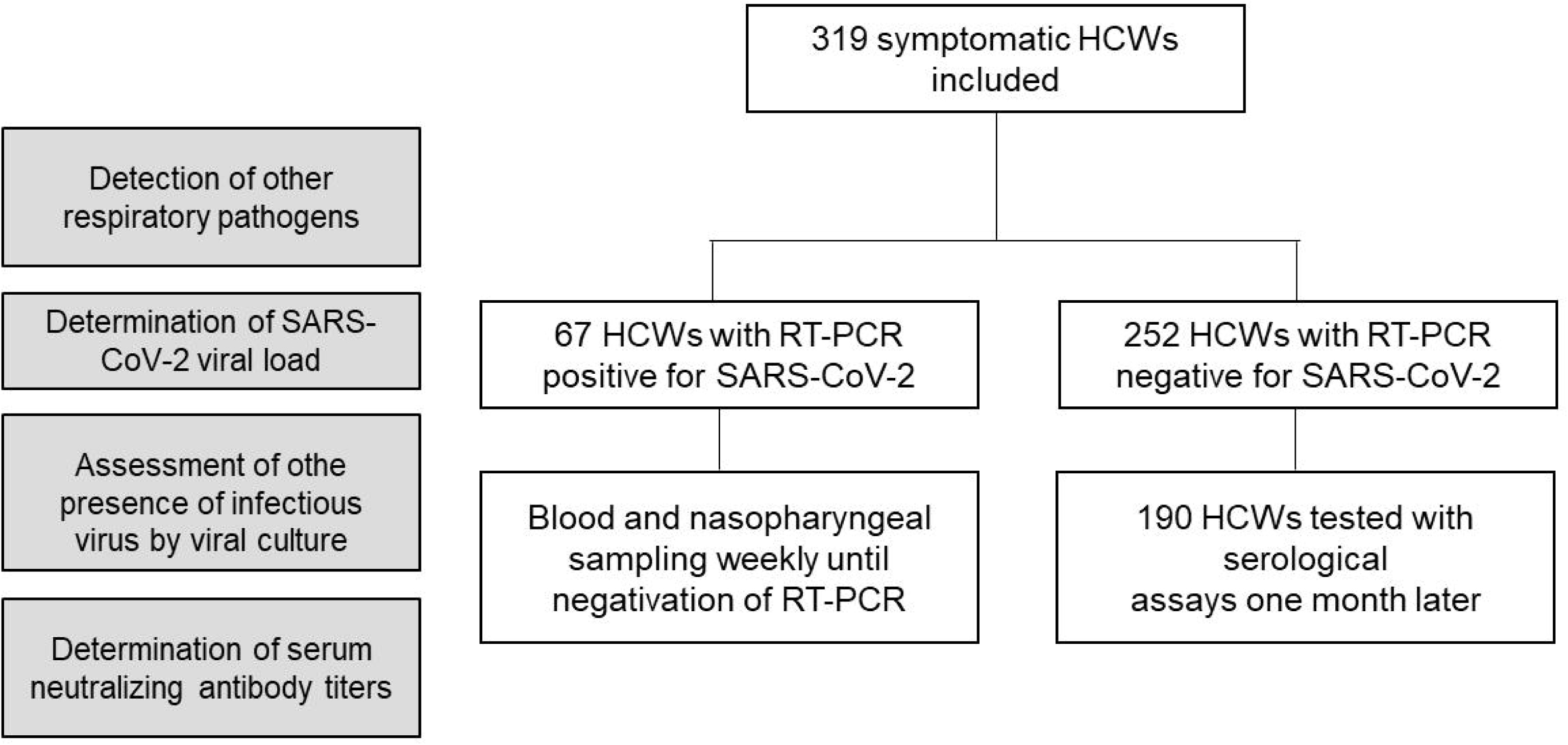
Flow diagram of study population – HCWs, healthcare workers.

### Microbiological investigations

HCWs were tested for SARS-CoV-2 with a real-time RT-PCR on NPS (Cobas® SARS-CoV-2 Test, Roche, Basel, Switzerland). NSP used were either in Copan Universal Transport Medium (UTM-RT®) or in Cobas® PCR medium tube.

To evaluate the number of symptomatic HCWs negative for SARS-COV-2 but positive for another pathogen and to evaluate the co-infection rate in COVID-19 HCWs, a total of 307 NPS collected at inclusion were tested with the BIOFIRE® Respiratory 2.1 *plus* Panel (RP2.1*plus*) detecting 23 respiratory pathogens including SARS-CoV-2 (bioMérieux, Lyon, France).

SARS-CoV-2 positive HCWs with a co-infection were removed from the rest of the analysis because symptoms could not be exclusively attributed to SARS-CoV-2.

For COVID-19 HCWs, the SARS-CoV-2 load was determined weekly from inclusion until becoming negative by RT-PCR using SARS-CoV-2 R-gene® kit (bioMérieux, Lyon, France).

Nucleic acid extraction was performed from 0.2 mL NPS on NUCLISENS® easyMAG® and amplification using Biorad CFX96. Quantitative viral load was determined using four internally developed quantification standards targeting the SARS-CoV-2 N gene: QS1 to QS4 respectively at 2.5.10^6^, 2.5.10^5^, 2.5.10^4^, 2.5.10^3^ copies/mL of a SARS-CoV-2 DNA standard. These QS were controlled and quantified using the Nanodrop spectrophotometer (ThermoFisher) and Applied Biosystems QuantStudio 3D Digital PCR.

In parallel, NPS were tested using the CELL Control R-GENE® kit (amplification of the HPRT1 housekeeping gene) that contains 2 quantification standards QS1 and QS2, respectively at 10^4^ copies/µL (50,000 cells/PCR i.e. 1.25.10^6^ cells/mL in our conditions) and 10^3^ copies/µL (5000 cells/PCR i.e. 1.25.10^5^ cells/mL in our conditions) of DNA standard, to normalize the viral load according to sampling quality.

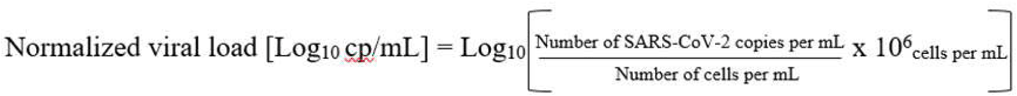

Viral culture was performed following interim biosafety guidelines established by WHO [29] from NPS in UTM-RT® only; guanidine contained in the Cobas PCR medium tube prevented culture assay due to cytotoxic activity. RT-PCR positive NPS were inoculated on confluent Vero cells (ATCC CCL-81) with Eagle’s Minimum Essential Media (EMEM) supplemented with 2% penicillin-streptomycin, 1% L-glutamine, and 2% inactivated fetal bovine serum. Plates were incubated at 33°C with 5% CO_2_ for 96 hours. Cytopathic effects (CPE) were monitored daily; samples were harvested when positive, while negative samples at 96 hours underwent subculture on new plates. Culture supernatants were sampled at 2 hours post-inoculation, at 96 hours, and at an additional 96 hours on subculture. RNA from supernatants was extracted by the automated MGISP-960 workstation using MGI Easy Magnetic Beads Virus DNA/RNA Extraction Kit (MGI Tech©, Marupe, Latvia), and SARS-CoV-2 detection was performed using TaqPath(tm) COVID-19 CE-IVD RT-PCR kit on a QuantStudio(tm) 5 System (Applied Biosystems, Thermo Fisher Scientific, Waltham, USA).

### Serological investigations

The presence of anti-SARS CoV-2 antibodies was evaluated on serum samples using the Wantai SARS-CoV-2 Ab ELISA kit (Wantai, Beijing, China), which detects total antibodies, and the VIDAS® SARS-COV-2 IgG test (bioMérieux, Lyon, France), according to the manufacturers’ instructions. Positivity was established according to the threshold value recommended by each manufacturer.

Neutralizing antibodies were quantified with a neutralization assay using lentiviral pseudotypes on serum samples. Briefly, gag/pol and luciferase plasmids were co-transfected with a SARS-CoV-2 full length S plasmid in HEK293T cells and pseudoviruses were harvested after 72h. Serial dilutions of human serum were incubated with pseudoviruses at 37°C for 1 h, then transferred onto HeLa-ACE2 cells in 96-well plates at 10 000 cells/well (Corning). Plates were incubated at 37°C for 48 hours and HeLa-ACE2 cells were further lysed using 1x luciferase lysis buffer (Oz Biosciences), at room temperature for 1h. Luciferase activity was measured by adding luciferase substrate (Oz Biosciences), according to the manufacturer’s instructions. Luciferase intensity was then read on a TECAN® luminometer. The results from this assay were expressed as the serum dilution required to reduce infection by 50% (neutralization titer).

### Statistical analysis

The median (interquartile range, IQR) was applied for describing continuous variables. The difference between groups was assessed by Student’s T test or Mann-Whitney U test, as appropriate. For categorical variables, n (%) was used for description and examined by Chi-square test or Fisher’s exact test. All statistical analyses were conducted by R (the R foundation, https://www.r-project.org/foundation/, version 3.6.1). Adjusted p-values were calculated using Benjamini & Hochberg method. An adjusted p-value <0.05 was considered statistically relevant.

### Ethics

The clinical study registered on ClinicalTrial.gov (NCT04341142) has been fully detailed [28]. Written informed consent was obtained from all participants and approval was obtained from the national review board for biomedical research in April 2020 (*Comité de Protection des Personnes Sud Méditerranée I*, Marseille, France; ID RCB 2020-A00932-37).

## Results

### Demographic and clinical characteristics of SARS-CoV-2 positive and negative HCWs

Between April 10th and May 28th, 2020, a total of 319 symptomatic HCWs were included (Figure 1). The main symptoms leading to SARS-CoV-2 screening were fever (61%, 194/319), cough (182/319, 57%) and asthenia (260/317, 82%). Among the 319 HCW, 67 (21.0%) were tested positive for SARS-CoV-2 (Table 1). The median [IQR] time between SARS-CoV-2 screening and symptom onset was 3 [2-6] and 5 [3-8] days for positive and negative patients, respectively (p=0.091). The median age of patients was 36 years old for positive and negative SARS-CoV-2 HCWs (p=0.93). Sex ratio (M/F) was 1:6 vs 1:4 for positive and negative patients respectively (p=0.64). The proportion of active smokers was lower in positive SARS-CoV-2 patients compared to negative ones (6/67 (8.9%) vs 64/252 (25.4%), p-adjusted=0.09). The proportion of smell and taste dysfunction was significantly higher in positive patients than in negative ones (26/67 (38.8%) vs 24/250 (9.5%) and 25/67 (37.3%) vs 27/250 (10.7%), respectively, p<0.001 for both symptoms). Diarrhea was reported in 23.9% (16/67) vs 41.3% (104/252) in positive and negative patients, respectively (p=0.091). Among the 67 positive patients, only two severe forms required conventional hospitalization, and one of them required ventilation support. No ICU admission was needed.

**Table 1.** Demographic and clinical characteristics of health-care workers exhibiting a negative or a positive RT-PCR test for SARS-CoV-2.*missing data

|  | Negative SARS-CoV-2<br>patients (n=252) | Positive SARS-CoV2<br>patients<br>(n= 67) | p-value | Adjusted p-value |
| --- | --- | --- | --- | --- |
| Gender. male. n (%) | 50 (19.84) | 9 (13.43) | 0.306 | 0.638 |
| Age. years. median [IQR] | 35.9 [27.5-47] | 36 [29.4-47.7] | 0.656 | 0.926 |
| Body Mass Index*. n | 249 | 67 | 0.148 | 0.473 |
| median [IQR] | 23.3 [21.1-27.1] | 24 [22.5-27.3] |  |  |
| Currently Smoker. n(%) | 64/252 (25.4) | 6/67 (8.96) | 0.013 | 0.091 |
| Alcohol consumption*. Daily. n(%) | 8/251 (3.19) | 3/67 (4.48) | 0.248 | 0.606 |
| Presence of Comorbidity*. n (%) | 109/249 (43.78) | 17/66 (25.76) | 0.012 | 0.091 |
| Description of Comorbidity. |  |  |  |  |
| Neurological disorders . n (%) | 7 (6.42) | 1 (5.88) | 1.000 | 1.000 |
| Cardiovascular disorders . n (%) | 2 (1.83) | 1 (5.88) | 0.355 | 0.649 |
| Hypertension. n (%) | 14 (12.84) | 3 (17.65) | 0.701 | 0.935 |
| Heart Failure. n (%) | 3 (2.75) | 1 (5.88) | 0.444 | 0.735 |
| Diabetes. n (%) | 5 (4.59) | 0 (0) | 1.000 | 1.000 |
| Immune deficiency . n (%) | 2 (1.83) | 0 (0) | 1.000 | 1.000 |
| Liver disease . n (%) | 2 (1.83) | 0 (0) | 1.000 | 1.000 |
| Kidney disease . n (%) | 1 (0.92) | 1 (5.88) | 0.253 | 0.606 |
| Cancer . n (%) | 2 (1.83) | 1 (5.88) | 0.355 | 0.649 |
| Hypothyroidie. n (%) | 11 (10.09) | 2 (11.76) | 0.688 | 0.935 |
| Rheumatic Disease. n (%) | 2 (1.83) | 2 (11.76) | 0.088 | 0.324 |
| Chronic respiratory disease . n (%) | 25 (22.94) | 1 (5.88) | 0.193 | 0.580 |
| HCW Contact patient. n(%) | 228/252 (90.48) | 64/67 (95.52) | 0.284 | 0.634 |
| Post-symptom delay. day. median [IQR] | 5 [3-8] | 3 [2-6] | 0.015 | 0.091 |
| Symptom. |  |  |  |  |
| Fever. n(%) | 149/252 (59.13) | 45/67 (67.16) | 0.291 | 0.634 |
| Sore throat*. n (%) | 55/250 (22) | 12/67 (17.91) | 0.576 | 0.837 |
| Diarrhoeas. n(%) | 104/252 (41.27) | 16/67 (23.88) | 0.014 | 0.091 |
| Pain. n(%) | 187/252 (74.21) | 53/67 (79.1) | 0.505 | 0.758 |
| Muscular. n(%) | 155 (82.89) | 50 (94.34) | 0.062 | 0.249 |
| Chest. n(%) | 38 (20.32) | 11 (20.75) | 1.000 | 1.000 |
| Joints. n(%) | 24 (12.83) | 3 (5.66) | 0.225 | 0.601 |
| Abdominal. n(%) | 72 (38.5) | 12 (22.64) | 0.048 | 0.232 |
| Asthenia*. n(%) | 204/250 (81.6) | 56/67 (83.58) | 0.845 | 1.000 |
| Rhinorrhea *. n(%) | 121/250 (48.4) | 46/67 (68.66) | 0.005 | 0.079 |
| Nauseas*. n(%) | 68/249 (27.31) | 14/67 (20.9) | 0.365 | 0.649 |
| Cough. n(%) | 135 (53.57) | 47 (70.15) | 0.022 | 0.115 |
| Shortness of breath*. n(%) | 96/249 (38.55) | 21/67 (31.34) | 0.346 | 0.649 |
| Headache. n(%) | 198 (78.57) | 54 (80.6) | 0.847 | 1.000 |
| Irritability*. n(%) | 65/250 (26) | 14/67 (20.9) | 0.485 | 0.758 |
| Anosmia. n(%) | 24 (9.52) | 26 (38.81) | <0.001 | <0.001 |
| Ageusia. n(%) | 27 (10.71) | 25 (37.31) | <0.001 | <0.001 |
| Ophthalmic pain. n(%) | 28 (11.11) | 6 (8.96) | 0.775 | 0.979 |
| Hospitalized*. n(%) | 0/179 (0) | 2/58 (3.45) | 0.059 | 0.249 |

\*missing data

### Investigation of bacterial and viral respiratory pathogens

To explore the potential presence of other respiratory infections in symptomatic HCWs tested negative for SARS-CoV-2 and to assess the co-infection rate in COVID-19 HCWs, a multiplex molecular respiratory panel was performed. The detection of SARS-CoV-2 with the multiplex panel was fully concordant with the initial routine diagnosis. Other respiratory pathogens were found in 6/66 (9.1%) vs 47/241 (19.5%) in SARS-CoV-2 positive and negative HCWs, respectively (p = 0.07) (Table 2). The pathogens responsible for co-infection in the 6 COVID-19 HCWs were rhinovirus/enterovirus (n=3), adenovirus (n=2), and parainfluenza virus 2 (n=2); one patient had a multiple infection (adenovirus - rhinovirus/enterovirus) in addition to SARS-CoV-2. Clinical and demographical characteristics of these 6 co-infected patients are detailed in supplementary Table 1. No bacterial co-infection was found for positive SARS-CoV-2 patients. For negative SARS-CoV-2 HCWs, the most frequent pathogens were rhinovirus/enterovirus (n=27), adenovirus (n=15) and other coronaviruses (HKU1 or NL63, n=7). *Chlamydia pneumoniae* (n=2) or *Mycoplasma pneumoniae* (n=1) were also found in negative SARS-CoV-2 HCWs (Table 2).

**Table 2.**
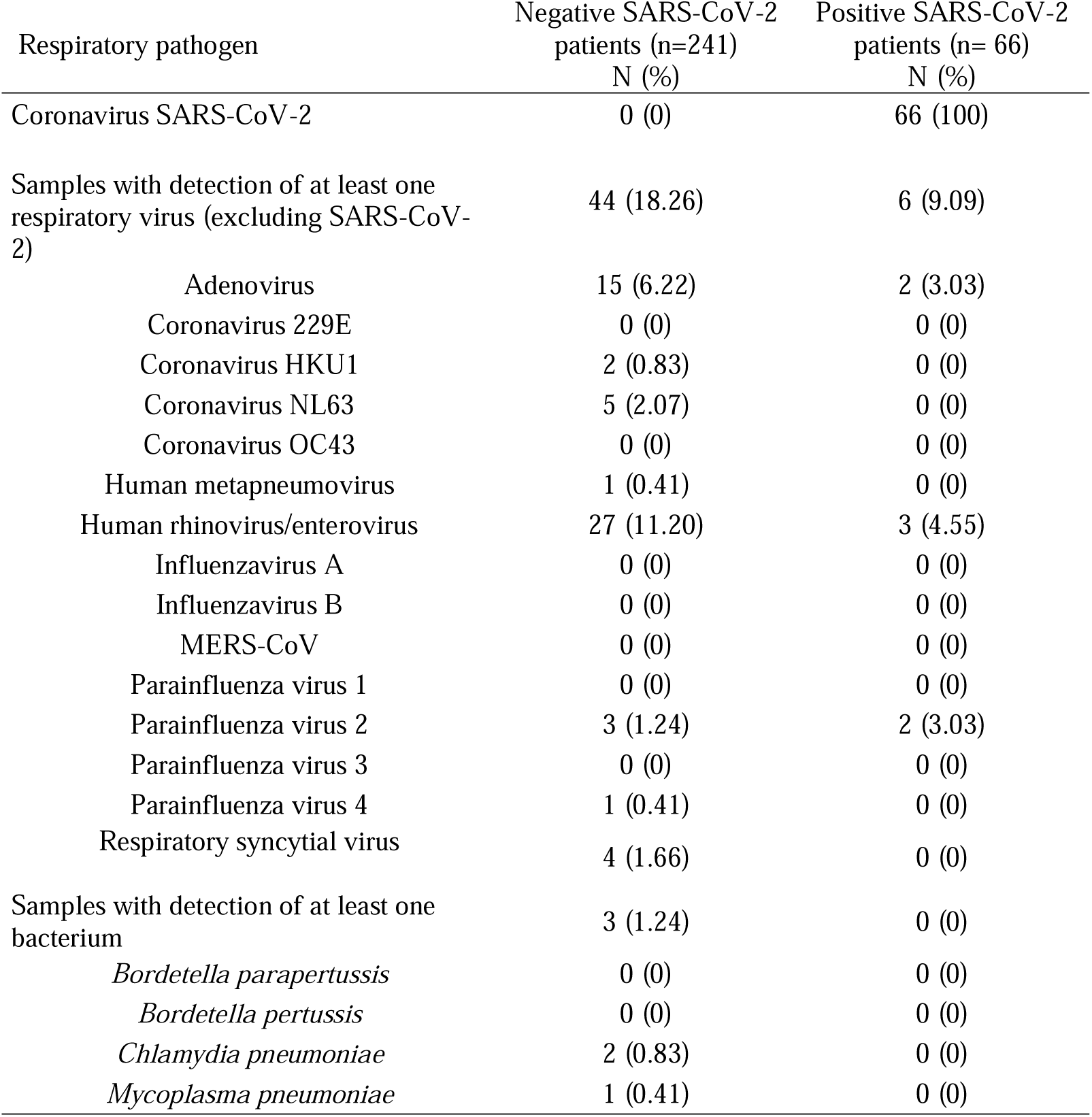
Investigation of respiratory pathogens in nasopharyngeal samples from symptomatic healthcare workers using the BIOFIRE® Respiratory Panel 2.1 plus. No significant statistical difference (p-value>0.05) was observed between samples tested negative or positive by SARS-CoV-2 RT-PCR.

| Respiratory pathogen | Negative SARS-CoV-2<br>patients (n=241)<br>N (%) | Positive SARS-CoV-2<br>patients (n= 66)<br>N (%) |
| --- | --- | --- |
| Coronavirus SARS-CoV-2 | 0 (0) | 66 (100) |
| Samples with detection of at least one<br>respiratory virus (excluding SARS-CoV-2) | 44 (18.26) | 6 (9.09) |
| Adenovirus | 15 (6.22) | 2 (3.03) |
| Coronavirus 229E | 0 (0) | 0 (0) |
| Coronavirus HKU1 | 2 (0.83) | 0 (0) |
| Coronavirus NL63 | 5 (2.07) | 0 (0) |
| Coronavirus OC43 | 0 (0) | 0 (0) |
| Human metapneumovirus | 1 (0.41) | 0 (0) |
| Human rhinovirus/enterovirus | 27 (11.20) | 3 (4.55) |
| Influenzavirus A | 0 (0) | 0 (0) |
| Influenzavirus B | 0 (0) | 0 (0) |
| MERS-CoV | 0 (0) | 0 (0) |
| Parainfluenza virus 1 | 0 (0) | 0 (0) |
| Parainfluenza virus 2 | 3 (1.24) | 2 (3.03) |
| Parainfluenza virus 3 | 0 (0) | 0 (0) |
| Parainfluenza virus 4 | 1 (0.41) | 0 (0) |
| Respiratory syncytial virus | 4 (1.66) | 0 (0) |
| Samples with detection of at least one<br>bacterium | 3 (1.24) | 0 (0) |
| *Bordetella parapertussis* | 0 (0) | 0 (0) |
| *Bordetella pertussis* | 0 (0) | 0 (0) |
| *Chlamydia pneumoniae* | 2 (0.83) | 0 (0) |
| *Mycoplasma pneumoniae* | 1 (0.41) | 0 (0) |

### Duration of SARS-CoV-2 PCR positivity according to normalized viral load

For the 61 SARS-CoV-2 positive HCWs without co-infection, normalized viral load was determined each week up to negativity of the RT-PCR test (Figure 2).

**Figure 2.**
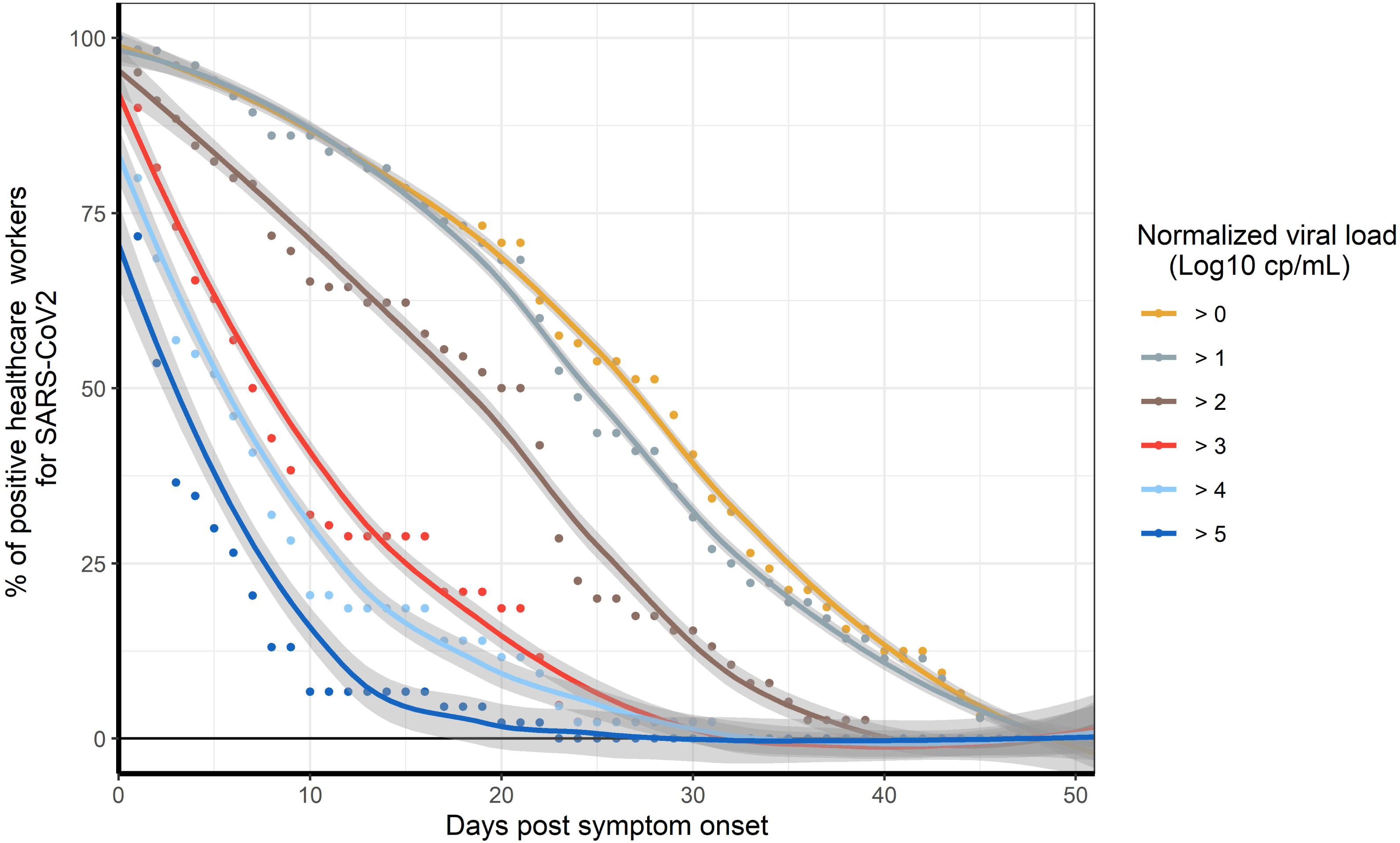
Longitudinal proportion of positive SARS-CoV-2 healthcare workers according to normalized viral load. Fit Loess curve represents local polynomial regression performed with Loess method. CI at 95% is indicated (grey area).

The percentage of HCWs with a viral load >5 log_10_ cp/mL, corresponding to a cycle threshold (Ct) value < 25, rapidly decreased during the first days after symptom onset and reached less than 15% at 8 days. At 20 days after symptom onset, about 40% of HCWs had a positive RT-PCR with a viral load > 2 log_10_ cp/mL. At 40 days post-symptom onset, SARS-CoV-2 RNA was still detectable for 12% of HCWs (< 1 log_10_ cp/mL, Ct value >37).

### Viral culture results according to viral load and date of symptom onset

A total of 64 SARS-CoV-2 RT-PCR positive NPS samples collected from 40 patients without co-infection were inoculated for cell culture (Figure 3A, supplementary Figure 1). Forty-two of these 64 samples (65.6%) showed a positive viral culture. The median [IQR] of normalized viral load for the culturable samples was 6.7 [5.6-7.4] log_10_ cp/mL vs 3.6 [2.9-4.9] log_10_ cp/ml for non-culturable specimens (p<0.001). The lowest viral load associated with culturable virus was 3.7 log_10_ cp/mL (Ct value of 30.2) (supplementary Figure 1, supplementary Table 2). More than 90% of samples (38/42) with culturable virus had a viral load > 4.5 log_10_ cp/mL, corresponding to a Ct value of 26 for the N gene, and were collected before 10 days after symptom onset.

**Figure 3.**
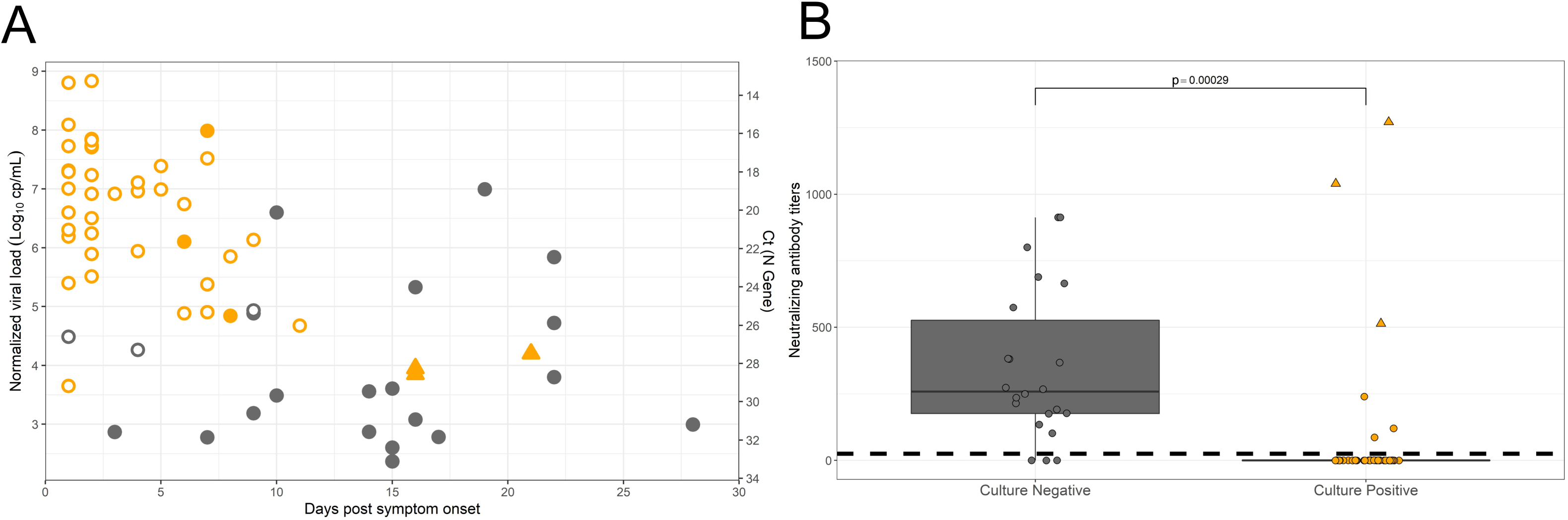
Viral culture results of SARS-CoV-2 according to post symptom delay and the presence of neutralizing antibodies. Black circles represent negative virus culture samples and orange circles or triangles represent the positive virus culture samples. Triangles correspond to samples positive on cell culture without cytopathic effect. Solid circles indicate samples with a presence of neutralizing antibodies while empty circles indicate the absence of neutralizing antibodies in serum. A. The Y-axis corresponds to the normalized viral load expressed in Log10 cp/mL or Cycle threshold (Ct) values. B. Dotted lines correspond to the limit of quantification of neutralizing antibody titers.

Further than twelve days after symptom onset, infectious virus could be retrieved in 3 samples, only after subculture. The positivity of these subcultures was established using an RT-PCR test performed on culture supernatant as the Ct difference was > 10 compared to the first passage despite the absence of CPE. These 3 samples had a viral load ranging from 3.85 to 4.20 log_10_ cp/mL and were collected from 2 patients. At 15 days after symptom onset for one patient, who presented a severe form, and at 16 and 21 days after symptom onset for the other with mild disease.

Neutralizing antibody (nAb) titers were measured in contemporaneous serum samples using a SARS-CoV-2 pseudo-typed virus assay. Among individuals with negative viral culture, a large fraction, 18/22, were positive for serum neutralizing activity, suggesting that nAbs may inhibit viral culture from respiratory samples. Of note, 6/7 individuals with a high viral load in NPS (>4.5 log_10_ cp/mL) and negative viral culture were positive for the detection of nAb. Conversely, high nAb titers were found in the three serum samples of the 2 HCWs with positive viral cultures more than twelve days after symptom onset. We also noticed that 3 additional NPS samples positive for viral culture were from individuals with a contemporaneous nAb activity but at very low titers (Figure 3B).

### Retrospective SARS-CoV-2 infection assessment with serological testing

Out of the 252 PCR negative HCW at initial screening, a total of 190 returned 1 month after inclusion (V5) for serological testing. Seven of 190 were seropositive at V5 with both of the selected serological assays. At inclusion, no other respiratory pathogens were detected with the multiplex respiratory panel for these seven individuals who developed mild respiratory symptoms (cough (5/7), shortness of breath (5/7) and rhinorrhea (3/7)). Interestingly, 4 out of the 7 suffered of ophthalmic pain, compared to 30 out of 270 in the rest of the cohort tested with the multiplex respiratory panel.

A problem in sampling quality did not appear to explain the negativity of the SARS-CoV-2 PCR from these 7 seropositive HCWs as the mean cell number in NPS from these individuals, as evaluated using a house keeping gene, was not significantly different from that of the rest of seronegative HCWs, at 4.0 10^4^ cells/PCR and 6.9 10^4^ cells/PCR, respectively (p-value = 0.14).

At inclusion, no anti-SARS-CoV-2 Abs had been detected for 6 of these seven HCWs. The one HCW who was already seropositive at inclusion was, however, included late in our study, at 24 days after symptom onset, which may explain the negativity of their PCR test. For the other 6 HCWs, the median (range) time between symptom onset and inclusion, i.e. RT-PCR screening, was 3 (1-4) days, as for PCR positive patients, suggesting that the negative SARS-CoV-2 RT-PCR cannot be explained by a delayed diagnosis. Taken together, SARS-CoV-2 detection was potentially missed in 6/190 (3.15%) HCWs by RT-PCR performed on nasopharyngeal swabs.

## Discussion

We analysed data obtained from front-line HCWs enrolled during the first wave of the COVID-19 pandemic. A high prevalence of COVID-19 was found, which is consistent with previous reports on this highly exposed population [7,30–32]. Regarding clinical findings, HCWs presented mostly mild disease, which can be explained by their relatively young age (median of 36 years) [33]. In addition, we confirmed the important proportion of smell and taste dysfunction in these mild COVID-19 patients. As already reported [7,34], the specificity of these symptoms in the present study was high (> 90%). We also found a low rate (9.1%) of co-infection in SARS-CoV-2 positive patients. Higher rates have been reported elsewhere which can be explained by several factors, including the severity of the disease and the timing of testing [35,36].

In the present study, only 3.2 % of HCWs with a negative RT-PCR test at inclusion developed a seroconversion one month later, suggesting the possibility of a false negative PCR test result at initial screening. Several factors can explain false negative results with RT-PCR, including the low sensitivity of some assays [37,38], a poor quality of the sample or an inappropriate timing of sampling [39]. However, the Cobas RT-PCR assay used for screening herein has been widely evaluated and a lack of sensitivity has not been reported [40]. Furthermore, the quality of the samples was checked with the use of a cell control [41] in order to prevent false negative results due to a lack of cells. Taken together, our findings confirmed a low rate (<5%) of false negative RT-PCR results using serology testing [42], although initial reports wrongly alerted the scientific community about a poor sensitivity of RT-PCR tests for COVID-19 screening [43]. Due to our study design, it cannot be excluded that some individuals may have developed COVID-19 between inclusion and the serological testing performed one month later for negative patients at inclusion.

Herein we found that a substantial (12%) part of the cohort was still RT-PCR positive 40 days post-symptom onset, which is consistent with a large study conducted in mild COVID-19 patients, in which 10% of patients had detectable RNA four weeks after symptom onset [14]. In a recent meta-analysis, a mean duration of RT-PCR positivity of 17 days has been reported with a maximum of 83 days [44]. It is important to emphasize that RT-PCR tests cannot distinguish between infectious virus and non-infectious RNA, and that RNA detection frequently outlasts the duration of infectivity. In the present study, we reported that upper respiratory samples from HCWs with positive RT-PCR ten days after symptom onset, even those with significant viral loads, were mainly (90%) found negative in viral culture.

Contagiousness is potentially dependent on many factors, including the presence of infectious virus and the presence of upper respiratory symptoms, but according to our results, contagiousness seems very unlikely 10 days after symptom onset in HCWs presenting a mild infection. These findings are consistent with others who reported a less than 6% probability of cultivating the virus after 10 [25] or 15 [20] days. Another large study on 3,790 SARS-CoV-2 positive samples inoculated for viral culture found that 1700/1941 (87.6%) of culturable samples were collected during the first week [45].

This delay might be in line with the time required for the elicitation of nAbs as suggested by the limited number of infectious virus found herein in samples with high viral load taken after seroconversion, and by the contemporaneous presence of nAbs in the serum of most individuals with negative viral culture. Similarly, in a report on hospitalized patients, serum nAb titers were independently associated with the absence of detection of infectious SARS-CoV-2. In addition, this study also found that some hospitalized patients with a low titer of neutralizing antibodies can still have a positive culture, as also observed in our cohort [20]. A viral load threshold could be useful to assess the presence of infectious virus. Herein, the median viral load of positive culture specimens was 6.67 log_10_ copies/mL. In a study among hospitalized patients, in samples with detectable infectious virus, the viral load median was8.14 log_10_ copies/mL [20], while in children another study reported a median of 7.2 log_10_ copies/mL [46].

In addition to age [17], the observed difference concerning the viral load threshold in predicting the presence of infectious virus might be related to the severity of the disease, as higher viral loads are usually noticed in severe patients [47].

Although the measured viral load can vary depending on the gene targeted by the quantitative RT-PCR, a viral load of 5 to 6 log_10_ copies/mL has been suggested as a proxy for the presence of infectious virus [18,22]. Our findings suggest a viral load threshold of 4,5 log10 copies/mL might be used in patients with mild disease. This viral load corresponds to a Ct value of 26 with the Argene RT-PCR kit targeting the N gene used in the present study. For most clinical laboratories that cannot afford true quantitative RT-PCR results, Ct values can therefore be used to assess the presence of infectious virus, as previously reported [22– 25,27,45]. However, as underlined by Han et al., Ct values are highly dependent on the RT-PCR used and can be affected by batch effect or PCR conditions. Therefore, Ct values should be interpreted with caution [48]. Combined with other management strategies [49], the duration of symptoms and the determination of viral load can be helpful to discontinue HCWs isolation, which should contribute to reduce the risk of staff shortage.

The present study has several limitations. First, the inclusion period between April to June corresponded to the second half of the first wave in France and led to a limited number of enrolled patients. Furthermore, the national lockdown may have had a substantial impact on the circulation of other respiratory viruses and the rate of co-infection is possibly underestimated. In addition, the date of symptom onset can be difficult to determine accurately. As asymptomatic HCWs were not screened, the prevalence of SARS-CoV-2 infection among HCWs was certainly underestimated in our cohort [4,7,32]. Finally, the results of viral culture must be considered with caution as this method can lack sensitivity, with its performance being highly dependent on the proper collection, transport, and rapidity of inoculation of samples on cells.

In conclusion, we confirm the high prevalence of SARS-CoV-2 infection among HCWs who developed mainly mild COVID-19. Our data suggest that true quantitative viral load measured by RT-PCR or, at least, Ct values can be useful for appreciating the infectiousness of infected HCWs. In addition, we show that these patients are unlikely infectious 10 days after symptom onset, despite a high viral load. Taken together, these data could be very helpful for defining rules for discontinuing isolation of HCWs and facilitating their safe return to work.

## Data Availability

All data are available in the manuscript

## Figure legends

**Supplementary Figure 1**. Normalized SARS-CoV-2 viral load according to viral culture results. The Y-axis corresponds to the normalized viral load expressed in Log10 cp/mL or Cycle threshold (Ct) values.

## Acknowledgements

We thank all the personnel of the occupational health and medicine department of Hospices Civils de Lyon who contributed to the sample collection. We thank Virginie Pitiot, the Clinical Research Associate, for her excellent work. We thank all the technicians from the virology laboratory whose work made it possible to obtain all these data. We thank Karima Brahima and all members of the clinical research and innovation department for their reactivity (DRCI, Hospices Civils de Lyon). We thank Come Barranger for his help regarding the implementation of the quantitative RT-PCR. And lastly, we thank all the health care workers for their participation in this clinical study.

## COVID-SER Study group

Adnot Jérôme, Alfaiate Dulce, Bal Antonin, Bergeret Alain, Boibieux André, Bonnet Florent, Bourgeois Gaëlle, Brunel-Dalmas Florence, Caire Eurydice, Charbotel Barbara, Chiarello Pierre, Cotte Laurent, d’Aubarede Constance, Durupt François, Escuret Vanessa, Fascia Pascal, Fassier Jean-Baptiste, Fontaine Juliette, Gaillot-Durand Lucie, Gaymard Alexandre, Gillet Myriam, Godinot Matthieu, Gueyffier François, Guibert Nicolas, Josset Laurence, Lahousse Matthieu, Lina Bruno, Lozano Hélène, Makhloufi Djamila, Massardier-Pilonchéry Amélie, Milon Marie-Paule, Moll Frédéric, Morfin Florence, Narbey David, Nazare Julie-Anne, Oria Fatima, Paul Adèle, Perry Marielle, Pitiot Virginie, Prudent Mélanie, Rabilloud Muriel, Samperiz Audrey, Schlienger Isabelle, Simon Chantal, Trabaud Mary-Anne, Trouillet-Assant Sophie

---

**Supplementary Table 1.** Demographic and clinical characteristics of SARS-CoV-2 positive health-care workers exhibiting a negative or a positive detection of at least another respiratory pathogen * missing data

|  | Positive SARS-CoV-2<br>patients (n= 61) | Positive SARS-CoV-2 patients<br>with co-infection (n= 6) | p-value | Adjusted p-value |
| --- | --- | --- | --- | --- |
| Gender. male. n (%) | 8 (13.11) | 1 (16.67) | 1.000 | 1.000 |
| Age. years. median [IQR] | 37.1 [29.4-47.9] | 31.5 [31.1-32.5] | 0.263 | 1.000 |
| Body Mass Index. median [IQR] | 24 [22.5-27.3] | 24.4 [22.8-25.2] | 0.974 | 1.000 |
| Current Smoker. n (%) | 6/61 (9.84) | 0/6 (0) | 0.785 | 1.000 |
| Alcohol consumption. Daily. n (%) | 3/61 (4.92) | 0/6 (0) | 1.000 | 1.000 |
| Presence of Comorbidity*. n (%) | 17/60 (28.33) | 0/6 (0) | 0.326 | 1.000 |
| Description of Comorbidity. |  |  |  |  |
| Neurological disorders n (%) | 1 (5.88) | 0 (0) |  |  |
| Cardiovascular disorders n (%) | 1 (5.88) | 0 (0) |  |  |
| Hypertension. n (%) | 3 (17.65) | 0 (0) |  |  |
| Heart Failure n (%) | 1 (5.88) | 0 (0) |  |  |
| Diabetes. n (%) | 0 (0) | 0 (0) |  |  |
| Immune deficiency n (%) | 0 (0) | 0 (0) |  |  |
| Liver disease. n (%) | 0 (0) | 0 (0) |  |  |
| Kidney disease. n (%) | 1 (5.88) | 0 (0) |  |  |
| Cancer. n (%) | 1 (5.88) | 0 (0) |  |  |
| Hypothyroidy. n (%) | 2 (11.76) | 0 (0) |  |  |
| Rheumatic Disease. n (%) | 2 (11.76) | 0 (0) |  |  |
| Chronic respiratory disease n (%) | 1 (5.88) | 0 (0) |  |  |
| HCW Contact patient. n (%) | 59/61 (96.72) | 5/6 (83.33) | 0.249 | 1.000 |
| Post-symptom delay. day. median [IQR] | 3 [2-6.2] | 4 [1.5-5] | 0.893 | 1.000 |
| Symptom. |  |  |  |  |
| Fever. n (%) | 41/61 (67.21) | 4/6 (66.67) | 1.000 | 1.000 |
| Sore throat. n (%) | 12/61 (19.67) | 0/6 (0) | 0.582 | 1.000 |
| Diarrhea. n (%) | 15/61 (24.59) | 1/6 (16.67) | 1.000 | 1.000 |
| Pain. n (%) | 48/61 (78.69) | 5/6 (83.33) | 1.000 | 1.000 |
| Muscular. n (%) | 45 (93.75) | 5 (100) | 1.000 | 1.000 |
| Chest. n (%) | 9 (18.75) | 2 (40) | 0.274 | 1.000 |
| Joints. n (%) | 3 (6.25) | 0 (0) | 1.000 | 1.000 |
| Abdominal. n (%) | 11 (22.92) | 1 (20) | 1.000 | 1.000 |
| Asthenia. n (%) | 51/61 (83.61) | 5/6 (83.33) | 1.000 | 1.000 |
| Rhinorrhea. n (%) | 41 (67.21) | 5 (83.33) | 0.657 | 1.000 |
| Nauseas. n (%) | 14 (22.95) | 0 (0) | 0.330 | 1.000 |
| Cough. n (%) | 44 (72.13) | 3 (50) | 0.353 | 1.000 |
| Shortness of breath. n (%) | 20 (32.79) | 1 (16.67) | 0.657 | 1.000 |
| Headache. n (%) | 49 (80.33) | 5 (83.33) | 1.000 | 1.000 |
| Irritability. n (%) | 12 (19.67) | 2 (33.33) | 0.597 | 1.000 |
| Anosmia. n (%) | 24 (39.34) | 2 (33.33) | 1.000 | 1.000 |
| Ageusia. n (%) | 24 (39.34) | 1 (16.67) | 0.399 | 1.000 |
| Ophthalmic pain. n (%) | 5 (8.2) | 1 (16.67) | 0.444 | 1.000 |
| Hospitalized. n (%) | 2/61 (3.85) | 0/6 (0) | 1.000 | 1.000 |

**Supplementary Table 2.** Normalized viral load and Ct-values according to viral culture results.

|  |  | Ct value ( N gene) |  |  | Log10 cp/mL |  |  |
| --- | --- | --- | --- | --- | --- | --- | --- |
|  | sample (n) | Median [IQR] | Highest | lowest | Median [IQR] | Highest | lowest |
| Culturable | n=42 | 19.46 [17.31-23.33] | 30.2 | 14.15 | 6.67 [5.6-7.37] | 8.84 | 3.65 |
| Non culturable | n=22 | 29.63 [25.77-30.99] | 34.48 | 18.61 | 3.58 [2.37-4.85] | 7 | 2.37 |

